# Progression Free Survival Prediction for Head and Neck Cancer using Deep Learning based on Clinical and PET-CT Imaging Data

**DOI:** 10.1101/2021.10.14.21264955

**Authors:** Mohamed A. Naser, Kareem A. Wahid, Abdallah S.R. Mohamed, Moamen Abobakr Abdelaal, Renjie He, Cem Dede, Lisanne V. van Dijk, Clifton D. Fuller

## Abstract

Determining progression-free survival (PFS) for head and neck squamous cell carcinoma (HNSCC) patients is a challenging but pertinent task that could help stratify patients for improved overall outcomes. PET/CT images provide a rich source of anatomical and metabolic data for potential clinical biomarkers that would inform treatment decisions and could help improve PFS. In this study, we participate in the 2021 HECKTOR Challenge to predict PFS in a large dataset of HNSCC PET/CT images using deep learning approaches. We develop a series of deep learning models based on the DenseNet architecture using a negative log-likelihood loss function that utilizes PET/CT images and clinical data as separate input channels to predict PFS in days. Internal model validation based on 10-fold cross-validation using the training data (N=224) yielded C-index values up to 0.622 (without) and 0.842 (with) censoring status considered in C-index computation, respectively. We then implemented model ensembling approaches based on the training data cross-validation folds to predict the PFS of the test set patients (N=101). External validation on the test set for the best ensembling method yielded a C-index value of 0.694. Our results are a promising example of how deep learning approaches can effectively utilize imaging and clinical data for medical outcome prediction in HNSCC, but further work in optimizing these processes is needed.

## 1 Introduction

Head and neck squamous cell carcinomas (HNSCC) are among the most prevalent cancers in the world. Approximately 890,000 new cases of HNSCC are diagnosed a year, and rates are projected to continue to increase [1]. While prognostic outcomes for HNSCC, particularly for oropharyngeal cancer (OPC), have improved over recent years, patients still have a significant probability of disease recurrence or death [2]. Determination of progression-free survival (PFS) in HNSCC is a highly challenging task since the ultimate healthcare outcomes of patients are driven by a complex interaction betweena large number of variables, including clinical demographics, treatment approaches, and underlying disease physiology. While risk prediction models based on clinical demographics for HNSCC have been developed in the past [3], these methods may lack prediction potential due to their use of a small number of simple variables or linear nature.

PET/CT imaging provides an avenue to combine anatomical with functional imaging to gauge the phenotypic properties of tumors. Distinct morphologic and metabolic information derived from PET/CT has been linked to the underlying pathology of HNSCC tumors and is invaluable in diagnosis, staging, and therapeutic assessment [4]. Therefore, PET/CT imaging data has been as an attractive biomarker in developing clinical outcome prediction models for HNSCC. Radiomic analysis of PET/CT images in HNSCC has been heavily investigated in the past, with models based on statistical, geometric, and texture information contained within regions of interest showing promise in predicting PFS [5, 6]. However, radiomic methods rely on hand-crafted features and pre-defined regions of interest, which can introduce bias into analysis pipelines [7]. Therefore, deep learning approaches, which do not require a-priori definition of regions of interest and where features are learned during the model training process, have been touted as attractive alternatives in the medical imaging domain [7–10]. However, while deep learning methods for clinical prediction are promising, theyare often bottlenecked by small homogenous training and evaluation datasets that limit the generalizability of models [11]. Therefore, developing and validating deep learning models for HNSCC outcome prediction on large multi-institutional datasets is of paramount importance.

The 2021 HECKTOR Challenge provides an opportunity to utilize large heterogenous training and testing PET/CT datasets with matched clinical data to further unlock the power of clinical prediction models for HNSCC. This manuscript describes the development and evaluation of a deep learning prediction model based on the DenseNet architecture that can implement PET/CT images and clinical data to predict PFS for HNSCC patients.

## 2 Methods

We developed and trained a deep learning model (2.3) through a cross validation procedure (2.4) for predicting the progression-free survival (PFS) endpoint of OPC patients (based on censoring status and time-to-event between PET/CT scan and event) using co-registered ^18^F-FDG PET and CT imaging data (2.1) and associated clinical data (2.2). The performance of the trained model for predicting PFS was validated using a internalcross-validation approach and applied to a previously unseen testing set (2.5).

### 2.1 Imaging Data

The data set used in this manuscript consists of co-registered ^18^F-FDG PET and CT scans for 325 OPC patients (224 patients used for training and 101 patients used for testing). All training and testing data were provided in Neuroimaging Informatics Technology Initiative (NIfTI) format and were released through AIcrowd [12] for the HECKTOR Challenge at MICCAI 2021 [13].

All images (i.e., PET and CT) were cropped to fixed bounding box volumes, provided with the imaging data by [12], of size 144×144×144 mm3 in the x, y and z dimensions and then resampled to a fixed image resolution of 1 mm in the x, y, and z dimensions using spline interpolation of order 3. The cropping and resampling codes used were based on the code provided by the HECKTOR Challenge organizers (https://github.com/voreille/hecktor). The CT intensities were truncated in the range of [-200, 200] Hounsfield Units (HU) to increase soft tissue contrast and then were normalized to a [-1, 1] scale. The intensities of PET images were normalized with z-score normalization. We used the Medical Open Network for AI (MONAI) [14] software transformation packages to rescale and normalize the intensities of the PET/CT images. Image processing steps used in this manuscript are displayed in **Figure 1A and 1B**.

**Fig. 1.**
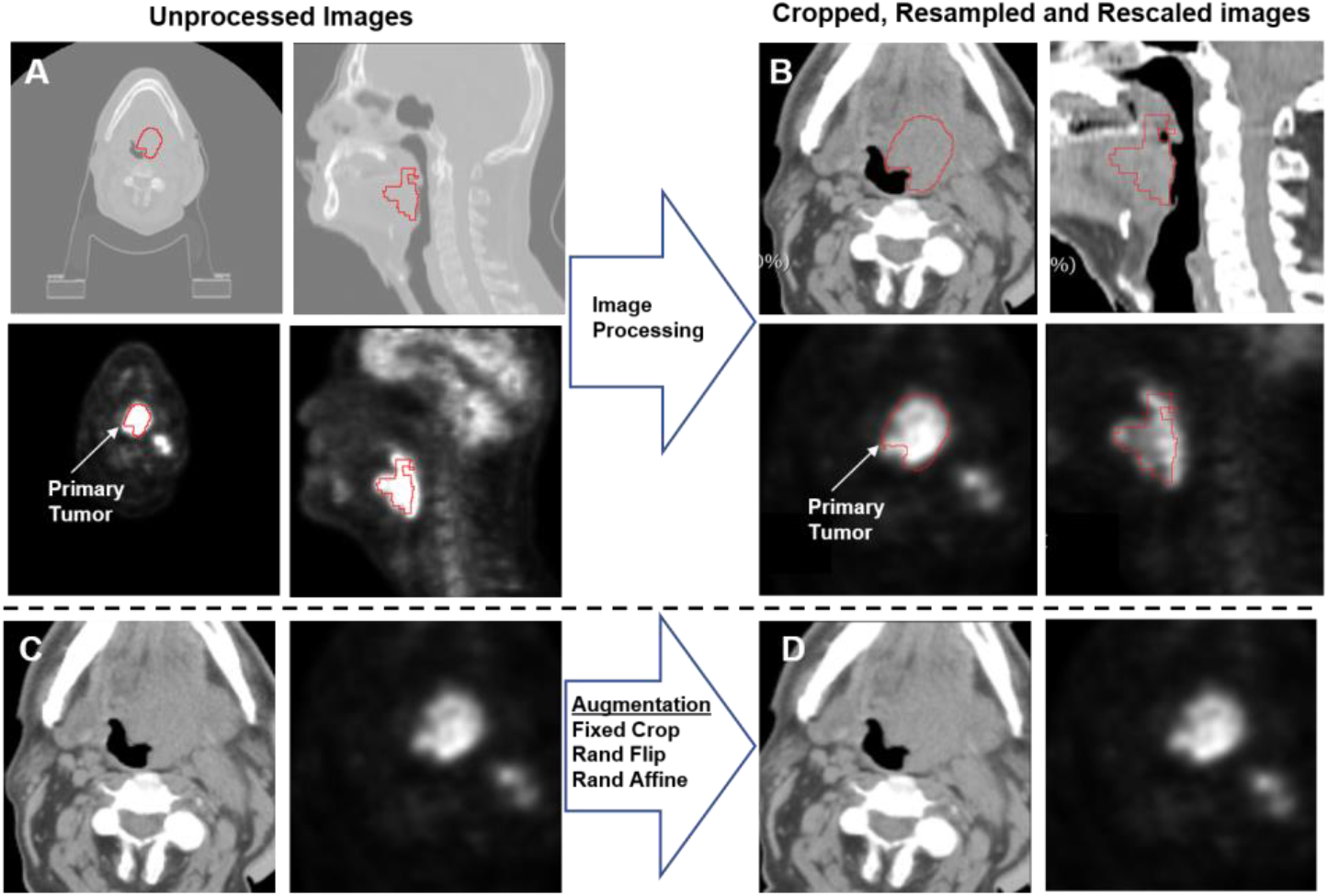
An illustration of the workflow used for image processing. (A) Overlays of the provided ground truth tumor segmentation mask (used to highlight tumor location in the image but not used in any analysis done in the paper) and the original CT (top) and PET (bottom) images. (B) Illustration of data augmentation that included cropping, random flips, and affine transformations used during the model training.

### 2.2 Clinical Data

Clinical data for patients were provided in .csv files, which included PFS outcome information and clinical variables. As per the HECKTOR website, “the progression is defined based on RECIST criteria: either a size increase of known lesions (change of T and or N), or appearance of new lesions (change of N and/or M). Disease-specific death is also considered a progression event for patients previously considered stable.”. We used all the clinical variables provided by the HECKTOR Challenge with complete fields (no NaN values present) for all patients. These variables included Center ID, Gender, Age, TNM edition, chemotherapy status, TNM group, T-stage, N-stage, and M-stage (9 variables in total). We elected not to use variables with incomplete fields (NaN values present), i.e., performance status, smoking status, etc., to avoid issues in the model building process. We mapped the individual T-, N-, and M-staging information to ordinal categoricalvariables in the following manner: T-stage: (‘Tx’: 0, ‘T1’: 1, ‘T2’: 2, ‘T3’: 3, ‘T4’: 4, ‘T4a’: 5, ‘T4b’: 6) – 7 classes; N-stage (‘N0’: 0, ‘N1’: 1, ‘N2’: ‘N2a’: 3, ‘N2b’: 4, ‘N2c’: 5, ‘N3’: 6) – 7 classes; M-stage (‘Mx’: 0, ‘M0’: 0, ‘M1’: 1) – 2 classes. We also mapped TNM group into an ordinal categorical variable based on information from the corresponding TNM stage in the following manner: (‘7 & I’: 0, ‘8 & I’: 0, ‘7 & II’: 1, ‘8 & II’: 1, ‘7 & III’: 2, ‘8 & III’: 2, ‘7 & IV’: 3, ‘8 & IV’: 3, ‘7 & IVA’: 4, ‘8 & IVA’: 4, ‘7 & IVB’: 5, ‘8 & IVB’: 5, ‘7 & IVC’: 6, ‘8 & IVC’: 6) – 7 classes. We then used a min-max rescaling approach provided by the scikit-learn Python package [15] for data normalization. The scaler was instantiated using only the training data and then applied to the test set, i.e., no data from the test set was allowed to leak into the training set. The normalized clinical data (9 variables per patient) was then reshaped such that each clinical variable is repeated 144×144×16 times in the x,y, and z dimensions, respectively and therefore a volumetric image of size 144×144×144 was formed from the 9 concatenated images of each clinical variable. The volumetric image generated from the clinical data was used as a third channel with the CT and PET images to the DenseNet model (2.3).

### 2.3 Model Architecture

A deep learning convolutional neural network model based on the DenseNet121 architecture included in the MONAI software package was used for the analysis. As shown in **Figure 2**, the network consists of 6, 12, 24, and 16 repetitions of dense blocks. Each dense block contained a pre-activation batch normalization, ReLU, and 3×3×3 convolution followed by a batch normalization, ReLU, and 1×1×1 convolution (DenseNet-B architecture). The network implementation in PyTorch used was provided by MONAI [14]. The model has 3 input channels with 144×144×144 each and 20 output channels representing different time intervals of the predicted survival probabilities (2.4).

**Fig. 2.**
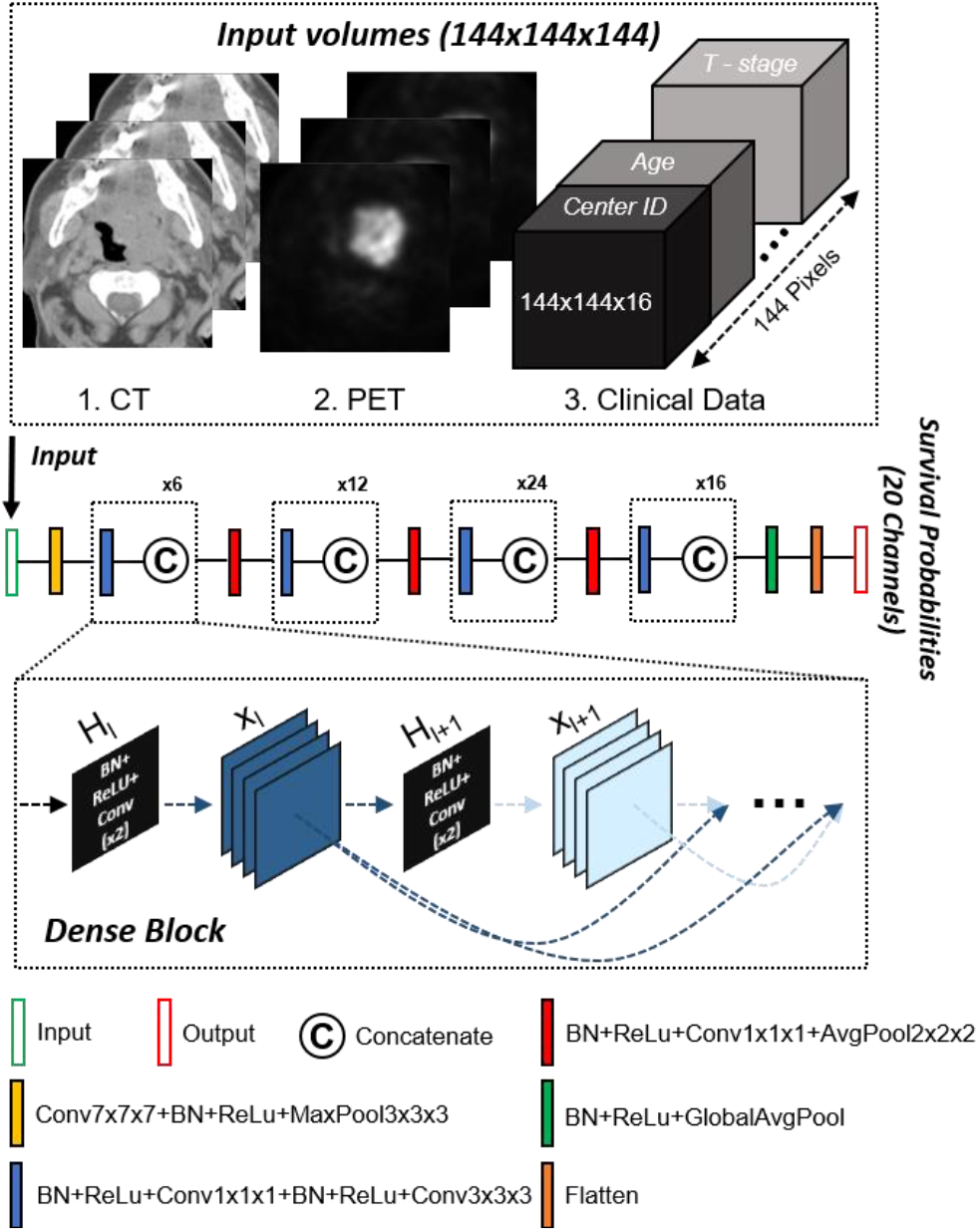
Schematic of the Densenet121 architecture used for the prediction model and the 3 input volumetric images representing CT, PET and clinical data. Clinical data is reformatted into a volumetric input for the model. The number of repeated dense blocks of (6, 12, 24, and 16) are given above each group of blocks.

### 2.4 Model Implementation

We used a 10-fold cross-validation approach where the 224 patients from the training data were divided into 10 non-overlapping sets. Each set (22 patients) was used for model validation while the remaining 202 patients in the remaining sets were used for training, i.e., each set was used once for testing and 9 times for training. The processed PET and CT (2.1) were cropped to a fixed size of (144, 144, 144) per image per patient. The clinical data was provided as a third channel to the model. We implemented additional data augmentation to the CT and PET channels, including randomhorizonal flips of 50% and random affine transformations with an axial rotation range of 12 degrees and a scale range of 10%. The image processing and data augmentation (2.1), network architecture (2.3) were used from the software packages provided by the MONAI framework [14]; code for these packages can be found at “https://github.com/Project-MONAI/”.

We implemented two main approaches for model building which utilizes imaging data only (Image), i.e., 2 channel input – CT/PET, or using imaging plus clinical data (Image+Clinical), i.e., 3 channel input – CT/PET + clinical data. We used a batch size of 2 patients’ images and clinical data and, therefore, the shape of the input tensor provided to the network (2.3) for the three-channel inputs was (2, 3, 144, 144, 144) and for the two-channel inputs was (2, 2, 144, 144, 144). The shape of the output tensor for 20 output channels was (2, 1, 20) for both Image and Image+Clinical models. The model was trained for 800 iterations with learning rate of (2×10^−4^ for iterations 0 to 300, 1×10^−4^ for iterations 301 to 600, and 5×10^−5^ for iterations 601 to 800). We used Adamas the optimizer and a minus log-likelihood function as the loss function [16]. An implementation example of the log-likelihood function compatible with Keras was provided in (https://github.com/chestrad/survival_prediction/blob/master/loss.py) by Kim et al. [16]. We modified the code to match with PyTorch tensors used by MONAI [14]. In our implementation, we divided the total time interval of 9 years, which covers all values reported for PFS in the training data set, into 20 discreate intervals of 164.25 days, representing the final 20 output channels of the network. The conditional probabilities of surviving in these intervals were obtained by applying a sigmoid function on the 20 outputs channels of the network. As shown in the illustrative example of **Fig. 3**, all the time intervals preceding the events were set to 1, and all other intervals were set to zero for both censored and uncensored patients (S vector). For uncensored patients (i.e., patients with censored status p = 1, where progression occurs), the time interval where progression occurs was set to 1 while all other intervals were set to 0 (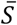 vector). The loss function is computed according to the equation shown in **Fig.3**.

**Fig. 3.**
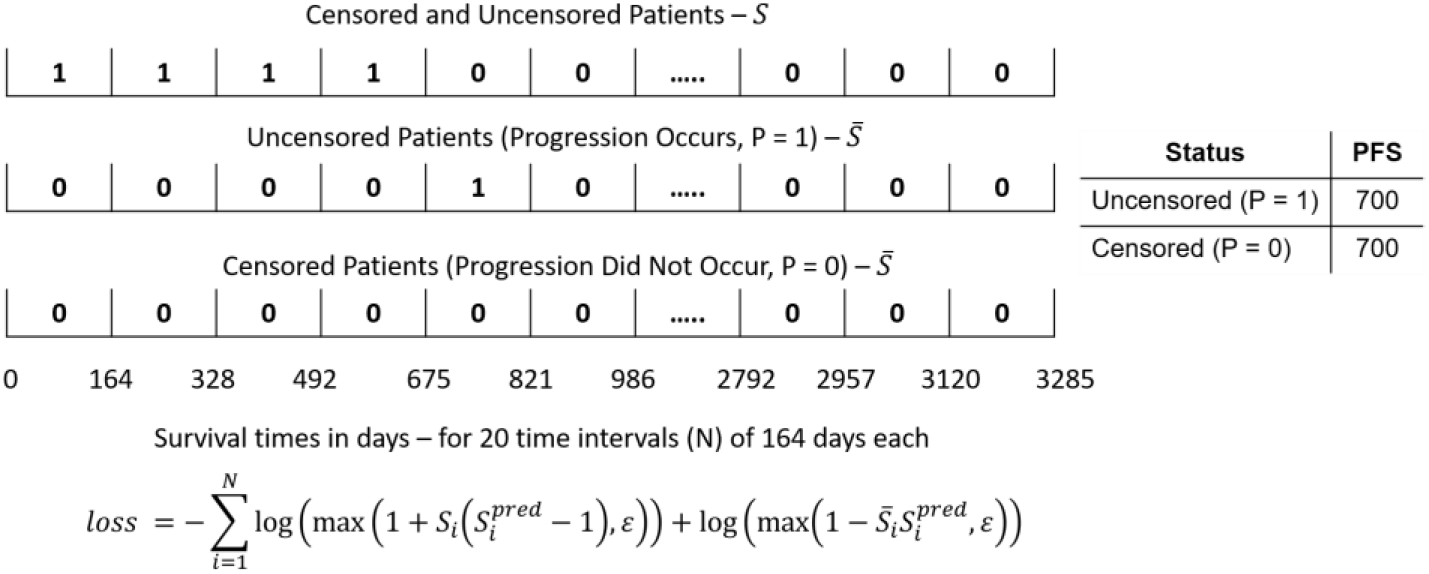
A graphical illustration of the −log likelihood loss function used by the DenseNet model.

We estimated the final PFS from the model predicted conditional probabilities by obtaining the summation of the cumulative probability of surviving each time interval times the duration of the time interval of 164.25 days. The cumulative probability of a time interval is obtained by multiplying the conditional probabilities of surviving all preceding time intervals times the time interval duration (i.e., 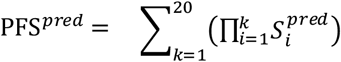 **\***(time interval duration = 164.25).

### 2.5 Model Validation

For each validation fold (i.e., 22 patients), we trained the DenseNet121 (2.3) on the remaining 202 patients. Therefore, we obtained 10 different trained models from 10-fold cross-validation. We evaluated the performance of each separate model on the corresponding validation set using concordance index (C-index) function of the lifelines package (https://github.com/CamDavidsonPilon/lifelines) which accounts for both censored and uncensored data through the indication of the time events occurrence of progression (i.e., progression occurs – True or False). We estimated the mean C-index by averaging all the C-index values obtained from each fold. It is also possible to measure the C-index by ignoring the events observed. Therefore, as an alternative metric, we also measured this modified C-index in reporting results.

For the test set (101 patients), we implemented two different model ensembling approaches to estimate the PFS. In the first approach, we estimated the PFS for each patient by each model and then obtained the mean value of the 10 predicted PFS values (AVERAGE approach). In the second approach, we first estimated the mean conditional probability survival vector by getting the mean value for each time interval. Then we computed the cumulative survival probability for each interval to estimate the consensus PFS values from the 10 models (CONSENSUS approach).

## 3 Results

The performance (predicted PFS predictions vs. ground truth) of each set of the 10-fold cross-validation procedure for the Image model and Image+Clinical model are shown in **Figure 4** and **Figure 5**, respectively. For both models, most individual fold predictions were not significantly different from the ground truth PFS (p>0.05), except sets 6 and 8 for the Image model and sets 2 and 6 for the Image+Clinical model. The cumulative mean performance measured over all folds for both models are shown in **Table 1**. Both models offered similar performance, regardless of the C-index calculation method.

**Table 1.** Mean C-index across all cross-validation folds for evaluated models (Image, Image+Clinical). C-index can be calculated with and without observed events; both calculations are reported.

| Model | C-index (Events used) | C-index (no Events used) |
| --- | --- | --- |
| Image | $0.842 \pm 0.080$ | $0.620 \pm 0.082$ |
| Image + Clinical | $0.841 \pm 0.045$ | $0.622 \pm 0.067$ |

**Fig. 4.**
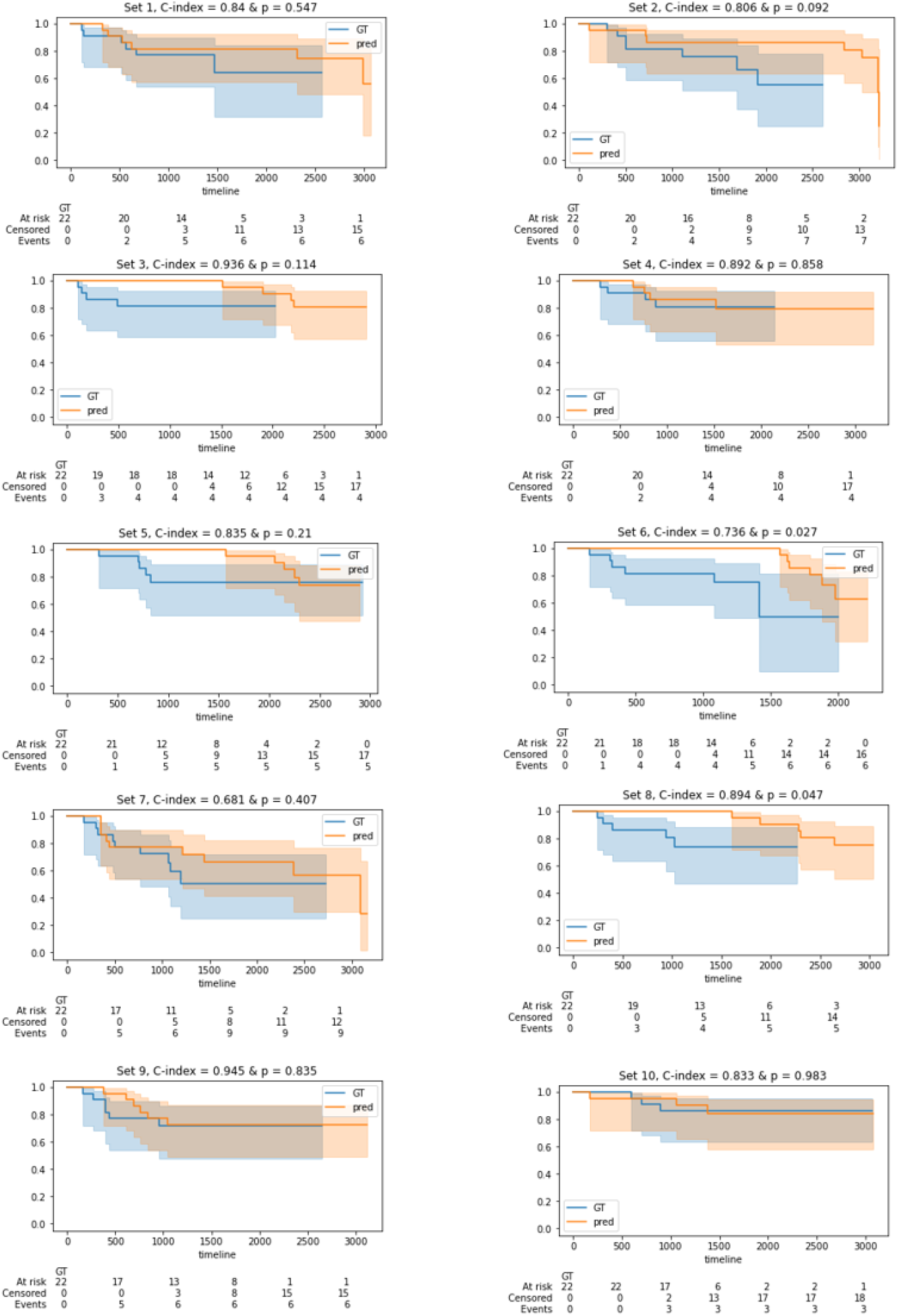
Kaplan Meier plots showing survival probabilities as a function of time in days for the ground truth (GT) PFS and the predicted PFS by the model using only imaging data (i.e., CT and PET) for each validation set of 22 patients. The C-index and the p-value of the logrank test for the GT and predicted PFS are shown above each subplot.

**Fig. 5.**
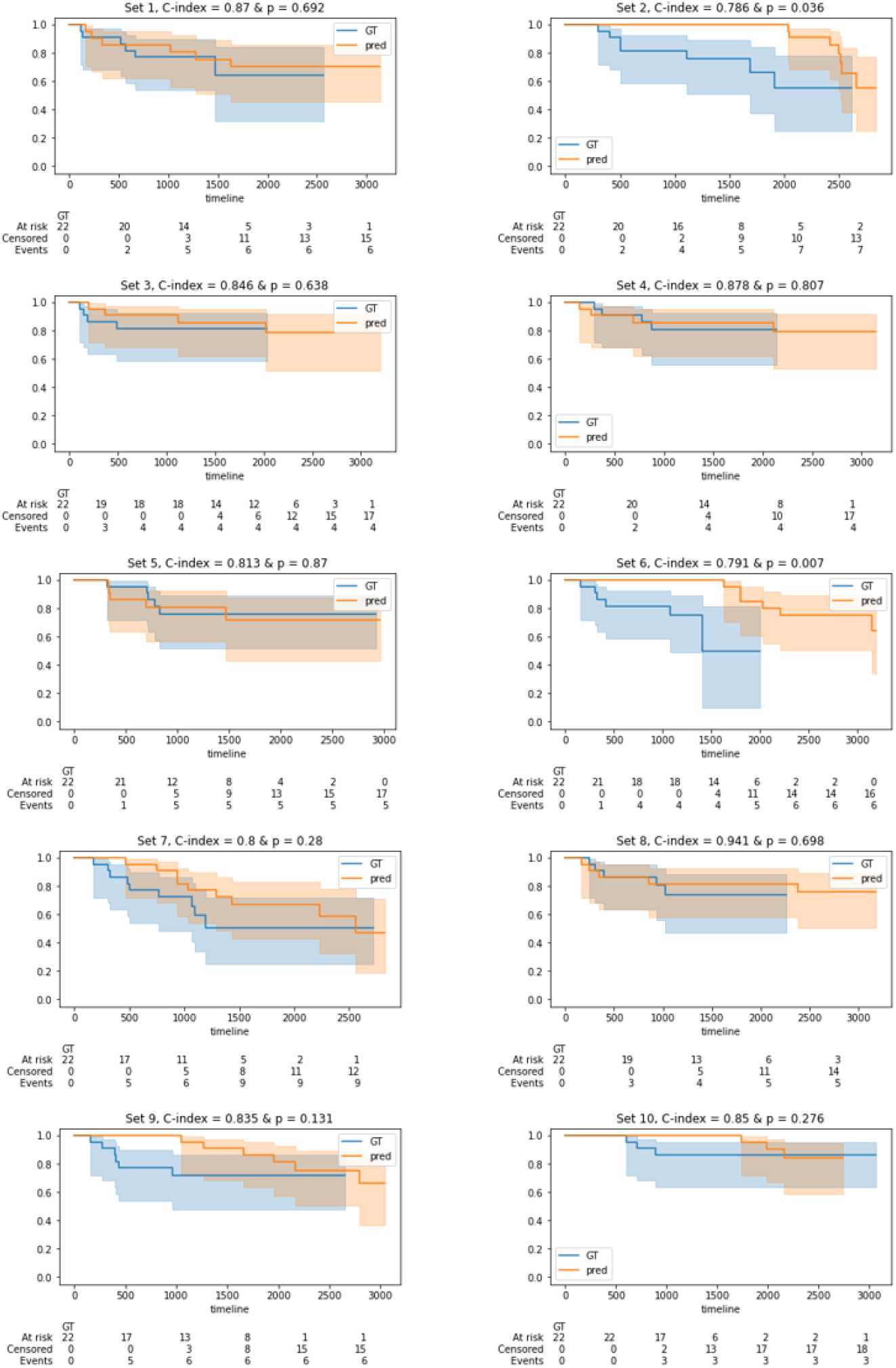
Kaplan Meier plots showing survival probabilities as a function of time in days for the ground truth (GT) PFS and the predicted PFS by the model using both imaging and clinical data for each validation set of 22 patients. The C-index and the p-value of the logrank test for the GT and predicted PFS are shown above each subplot.

When evaluated through external validation (test set), our model predictions fromeach cross-validation fold were ensembled using two methods (AVERAGE and CONSENSUS), leading to a total of four tested models (Image AVERAGE, Image CONSENSUS, Image+Clincial AVERAGE, Image+Clinical CONSENSUS). The results on the test set for these separate models are shown in **Table 2**. The highest performing model overall was the Image+Clinical model with a C-index of 0.694.

**Table 2.** Progression-free survival prediction measured using the C-index for ensemble models submitted to the 2021 HECKTOR Challenge evaluation portal. Two ensemble approaches were tested (AVERAGE, CONSENSUS). * The exact method of C-index calculation was not provided but can be inferred to be with no events used.

| Model | C-index* |
| --- | --- |
| Image AVERAGE | 0.645 |
| Image + Clinical AVERAGE | 0.689 |
| Image CONSENSUS | 0.651 |
| Image + Clinical CONSENSUS | 0.694 |

## 4 Discussion

In this study, we have utilized deep learning approaches based on DenseNet applied to PET/CT images and clinical data of HNSCC patients to predict PFS. The determination of prognostic outcomes is an unmet need for HNSCC patients that could improve clinical decision-making processes. While the performance of our models (as measured in C-index without events observed) is not ideal, they are reasonable within the context of prognostic prediction, which is known to be notoriously complextask [6]. Our main innovation stems from the use of ensembling approaches applied to predictions from internal cross-validation, which seems to reasonably improve overall performance on unseen data.

We evaluated two approaches (PET/CT inputs with and without clinical data) for internal validation through a 10-fold cross-validation approach in the training set to gauge performance before applying the models to the test set. When investigating individual cross-validation sets, we find that performance for both models is often comparable to the ground truth PFS. Specifically, when utilizing C-index with observed events information considered, models could reach fairly high prediction performance, with values up to 0.842 for the Image model. Compared to PFS prediction models evaluated with the C-indexin other studies, this is notably higher [5]. However, when implementing C-index calculations without considering observed events information, this value drops precipitously for both models. Specifically, a maximum value of 0.622 is achieved, which is more consistent with previously reported values for similar tasks. Interestingly, the addition of clinical data generally did not make any noticeable improvements in model performance, regardless of the C-index metric calculation method, possibly indicating the majority of informative data was contained within the PET/CT images. This observation runs counter to results seen in other clinical prediction models, where the addition of clinical data to imaging information often improves performance [6].

For the external validation (testing set) we further investigate the effect of specific model ensembling techniques through the AVERAGE and CONSENSUS methods. Generally, the CONSENSUS method (consensus from cumulative survival probability derived from mean conditional probability survival vectors) seems to offer per-formance gains over the AVERAGE method. Importantly, model performance on the test set is improved compared to the training set (assuming C-indexcalculation did not incorporate the observed events). Specifically, we achieve C-index gains of approximately 0.07 for test set performance. This may be due to the ensemble approach adding generalizability acquired from the combined inference capabilities shared across multiple cross-validation sets. Interestingly, while adding clinical data did not make noticeable differences in the training set, this was not the case in the test set, which offered substantial C-indexgains of approximately 0.04. The discrepancy may be explained by clinical data being more relevant when used in conjunction with model ensembling. Model ensembling is known to be a powerful technique in machine learning generally. This has been reiterated in other tasks for the HECKTOR Challenge, where ensembling provided impressive performance gains for tumor segmentation [17]. Therefore, we emphasize that model ensembling techniques may also be relevant for deep learning prediction models in HNSCC.

While we have taken steps to ensure a robust analysis, our study contains several limitations. Firstly, while we have included clinical data in our model-building process, we have not included all the data initially provided by the HECKTOR Challenge. Specifically, we have not included clinical data with incomplete fields (NaNs) for anypatients, such as tobaccostatus, alcohol status, and human papillomavirus status. Importantly, it is precisely these variables that are often the most highly correlated to prognosis in HNSCC [2, 18]. Therefore, data imputation techniques or related methods should be implemented in future studies to fully realize available clinical variables’ discriminative capabilities. A second limitation of our approach is we have used an out-of-the-box DenseNet architecture that has not been specifically optimized for imaging data combined with tabular clinical data. Moreover, to ensure that we could utilize our DenseNet architecture with an additional channel input for clinical data, we have concatenated clinical data into a 3D volume for model input. While using an open-source commonly available approach improves reproducibility, further studies should determine how architectural modifications can be made to optimize performance on this specific task. Additionally, in our model design process, we have selected discrete intervals based on observations in the training data. However, this assumes the training data fully encapsulates the PFS landscape, which may not necessarily be the case. Finally, since deep learning ensembling applied to PFS prediction is relatively understudied, we have approached model ensembling of individual cross-validation folds through relatively simple methods that involve linear combinations of individual model predictions. More complexmethods for deep learning ensembling [19] may provide additional predictive power for PFS tasks.

## 5 Conclusion

Using PET/CT and clinical data inputs, we developed, trained, and validated a series of deep learning models that could predict PFS in HNSCC patients in large-scale datasets provided by the 2021 HECKTOR Challenge. Cross-validation performance on the training set achieved mean C-index values of up to 0.622 (0.842 with alternative C-index calculations). Simple model ensembling approaches improved this performance further with reported C-index values up to 0.694 on the testing set. While our models offer modest performance on test data, these methods can be enhanced through additional clinical inputs, improved architectural modifications, and alternative ensembling approaches.

## Data Availability

Data are available from the MICCAI HECKTOR 2021 Challenge Organizers.

## Acknowledgements

M.A.N. is supported by a National Institutes of Health (NIH) Grant (R01 DE028290-01). K.A.W. is supported by a training fellowship from The University of Texas Health Science Center at Houston Center for Clinical and Translational Sciences TL1 Program (TL1TR003169), the American Legion Auxiliary Fellowship in Cancer Research, and a NIDCR F31 fellowship (1 F31 DE031502-01). C.D.F. received funding from the National Institute for Dental and Craniofacial Research Award (1R01DE025248-01/R56DE025248) and Academic-Industrial Partnership Award (R01 DE028290), the National Science Foundation (NSF), Division of Mathematical Sciences, Joint NIH/NSF Initiative on Quantitative Approaches to Biomedical Big Data (QuBBD) Grant (NSF 1557679), the NIH Big Data to Knowledge (BD2K) Program of the National Cancer Institute (NCI) Early Stage Development of Technologies in Biomedical Computing, Informatics, and Big Data Science Award (1R01CA214825), the NCI Early Phase Clinical Trials in Imaging and Image-Guided Interventions Program (1R01CA218148), the NIH/NCI Cancer Center Support Grant (CCSG) Pilot Research Program Award from the UT MD Anderson CCSG Radiation Oncology and Cancer Imaging Program (P30CA016672), the NIH/NCI Head and Neck Specialized Programs of Research Excellence (SPORE) Developmental Research Program Award (P50 CA097007) and the National Institute of Biomedical Imaging and Bioengineering (NIBIB) Research Education Program (R25EB025787). He has received direct industry grant support, speaking honoraria and travel funding from Elekta AB.

## Notes

### Competing Interest Statement

The authors have declared no competing interest.

### Author Declarations

All data was used with approval from the HECKTOR Challenge organizers. Institutional Review Boards (IRB) of all participating institutions permitted the use of images and clinical data, either fully anonymized or coded, from all cases for research purposes, only. Retrospective analyses were performed following the relevant guidelines and regulations as approved by the respective institutional ethical committees with protocol numbers: MM-JGH-CR15-50 (HGJ, CHUS, HMR, CHUM) and CER-VD 2018-01513 (CHUV).

## References

1. Johnson, D.E., Burtness, B., Leemans, C.R., Lui, V.W.Y., Bauman, J.E., Grandis, J.R.: Head and neck squamous cell carcinoma. Nat. Rev. Dis. Prim. 6, 1 –22 (2020).

2. Chow, L.Q.M.: Head and neck cancer. N. Engl. J. Med. 382, 60–72 (2020).

3. Budach, V., Tinhofer, I.: Novel prognostic clinical factors and biomarkers for outcome prediction in head and neck cancer: a systematic review. Lancet Oncol. 20, e313–e326 (2019).

4. Goel, R., Moore, W., Sumer, B., Khan, S., Sher, D., Subramaniam, R.M.: Clinical practice in PET/CT for the management of head and neck squamous cell cancer. Am. J. Roentgenol. 209, 289–303 (2017).

5. Haider, S.P., Burtness, B., Yarbrough, W.G., Payabvash, S.: Applications of radiomics in precision diagnosis, prognostication and treatment planning of head and neck squamous cell carcinomas. Cancers head neck. 5, 1 –19 (2020).

6. Chinnery, T., Arifin, A., Tay, K.Y., Leung, A., Nichols, A.C., Palma, D.A., Mattonen, S.A., Lang, P.: Utilizing Artificial Intelligence for Head and Neck Cancer Outcomes Prediction From Imaging. Can. Assoc. Radiol. J. 72, 73 –85 (2021).

7. Hosny, A., Aerts, H.J., Mak, R.H.: Handcrafted versus deep learning radiomics for prediction of cancer therapy response. Lancet Digit. Heal. 1, e106–e107 (2019).

8. Sun, Q., Lin, X., Zhao, Y., Li, L., Yan, K., Liang, D., Sun, D., Li, Z.-C.: Deep learning vs. radiomics for predicting axillary lymph node metastasis of breast cancer using ultrasound images: don’t forget the peritumoral region. Front. Oncol. 10, 53 (2020).

9. Avanzo, M., Wei, L., Stancanello, J., Vallières, M., Rao, A., Morin, O., Mattonen, S.A., El Naqa, I.: Machine and deep learning methods for radiomics. Med. Phys. 47, e185– e202 (2020).

10. Hosny, A., Parmar, C., Coroller, T.P., Grossmann, P., Zeleznik, R., Kumar, A., Bussink, J., Gillies, R.J., Mak, R.H., Aerts, H.J.W.L.: Deep learning for lung cancer prognostication: a retrospective multi-cohort radiomics study. PLoS Med. 15, e1002711 (2018).

11. Willemink, M.J., Koszek, W.A., Hardell, C., Wu, J., Fleischmann, D., Harvey, H., Folio, L.R., Summers, R.M., Rubin, D.L., Lungren, M.P.: Preparing medical imaging data for machine learning. Radiology. 295, 4–15 (2020).

12. AIcrowd MICCAI 2020: HECKTOR Challenges.

13. Andrearczyk, V., Valentin, O., Mario, J., Vallières, M., Castelli, J., Elhalawani, H., Boughdad, S., Prior, J.O., Depeursinge, A.: Overview of the HECKTOR challenge at MICCAI 2020: Automatic Head and Neck Tumor Segmentation in PET/CT.

14. Project MONAI.

15. Pedregosa, F., Varoquaux, G., Gramfort, A., Michel, V., Thirion, B., Grisel, O., Blondel, M., Prettenhofer, P., Weiss, R., Dubourg, V.: Scikit - learn: Machine learning in Python. J. Mach. Learn. Res. 12, 2825–2830 (2011).

16. Kim, H., Goo, J.M., Lee, K.H., Kim, Y.T., Park, C.M.: Preoperative CT-based Deep Learning Model for Predicting Disease-Free Survival in Patients with Lung Adenocarcinomas. https://doi.org/10.1148/radiol.2020192764. 296, 216–224 (2020). https://doi.org/10.1148/RADIOL.2020192764.

17. Iantsen, A., Visvikis, D., Hatt, M.: Squeeze-and-excitation normalization for automated delineation of head and neck primary tumors in combined PET and CT images. In: 3D Head and Neck Tumor Segmentation in PET/CT Challenge. pp. 37 –43. Springer (2020).

18. Leemans, C.R., Snijders, P.J.F., Brakenhoff, R.H.: The molecular landscape of head and neck cancer. Nat. Rev. Cancer. 18, 269 –282 (2018).

19. Ganaie, M.A., Hu, M.: Ensemble deep learning: A review. arXiv Prepr. 2104.02395. (2021).

